# Assessing accuracy and legitimacy of multimodal large language models on Japan Diagnostic Radiology Board Examination

**DOI:** 10.1101/2025.06.23.25329534

**Authors:** Yuichiro Hirano, Soichiro Miki, Yosuke Yamagishi, Shouhei Hanaoka, Takahiro Nakao, Tomohiro Kikuchi, Yuta Nakamura, Yukihiro Nomura, Takeharu Yoshikawa, Osamu Abe

**Author notes:** Corresponding author: Yuichiro Hirano.

## Abstract

**Purpose:** To assess and compare the accuracy and legitimacy of multimodal large language models (LLMs) on the Japan Diagnostic Radiology Board Examination (JDRBE).

**Materials and methods:** The dataset comprised questions from JDRBE 2021, 2023, and 2024, with ground-truth answers established through consensus among multiple board-certified diagnostic radiologists. Questions without associated images and those lacking unanimous agreement on answers were excluded. Eight LLMs were evaluated: GPT-4 Turbo, GPT-4o, GPT-4.5, GPT-4.1, o3, o4-mini, Claude 3.7 Sonnet, and Gemini 2.5 Pro. Each model was evaluated under two conditions: with inputting images (vision) and without (text-only). Performance differences between the conditions were assessed using McNemar’s exact test. Two diagnostic radiologists (with 2 and 18 years of experience) independently rated the legitimacy of responses from four models (GPT-4 Turbo, Claude 3.7 Sonnet, o3, and Gemini 2.5 Pro) using a five-point Likert scale, blinded to model identity.

Legitimacy scores were analyzed using Friedman’s test, followed by pairwise Wilcoxon signed-rank tests with Holm correction.

**Results:** The dataset included 233 questions. Under the vision condition, o3 achieved the highest accuracy at 72%, followed by o4-mini (70%) and Gemini 2.5 Pro (70%). Under the text-only condition, o3 topped the list with an accuracy of 67%. Addition of image input significantly improved the accuracy of two models (Gemini 2.5 Pro and GPT-4.5), but not the others. Both o3 and Gemini 2.5 Pro received significantly higher legitimacy scores than GPT-4 Turbo and Claude 3.7 Sonnet from both raters.

**Conclusion:** Recent multimodal LLMs, particularly o3 and Gemini 2.5 Pro, have demonstrated remarkable progress on JDRBE questions, reflecting their rapid evolution in diagnostic radiology.

Secondary abstract

Eight multimodal large language models were evaluated on the Japan Diagnostic Radiology Board Examination. OpenAI’s o3 and Google DeepMind’s Gemini 2.5 Pro achieved high accuracy rates (72% and 70%) and received good legitimacy scores from human raters, demonstrating steady progress.

## Introduction

Large language models (LLMs) possess rich medical knowledge. Studies have shown that recent LLMs can pass medical licensing examinations in multiple countries with scores far exceeding those of average human examinees [1–3]. LLMs have acquired multimodal capabilities, accepting images, video, and audio as input. Researchers have investigated the vision capabilities of multimodal LLMs, also known as vision-language models or VLMs, in various medical domains.

So far, the “eyes” of publicly available multimodal LLMs have not proven very impressive in the field of diagnostic radiology. Our previous research showed that GPT-4 Turbo with Vision, one of the earliest multimodal LLMs, did not outperform its text-only counterpart on the Japan Diagnostic Radiology Board Examination (JDRBE) [4]. Furthermore, due to the model’s poor image interpretation capability, the addition of images resulted in significantly worse scores in subjective rating by radiologists.

Likewise, GPT-4o, Gemini Flash 1.5, Gemini Pro 1.5, and Claude 3 Opus failed to benefit from the addition of images [5,6]. Similar findings have been reported for the Japanese Nuclear Medicine Board Examination [7]. Meanwhile, Kurokawa et al. found that diagnostic performance of Claude 3 Opus and Claude 3.5 Sonnet significantly improved when images were provided, using cases from *Radiology*’s “Diagnosis Please” [8]. Others have investigated the performance of multimodal LLMs on Japanese [9], European [10], and Australian [11] board certification examinations for diagnostic radiology, although they did not directly compare performance with and without images.

For now, the strength of “multimodal” LLMs appears to lie largely in their robust text-based knowledge rather than their ability to directly interpret medical images. Even if their scores are high in an image-rich examination like JDRBE, it cannot be definitively concluded that they have exceeded the image interpretation abilities of human radiologists. Furthermore, the ability of multimodal LLMs to provide diagnostic reasoning based on image interpretation has been insufficiently evaluated and remains undemonstrated in prior studies.

In early 2025, multiple new multimodal LLMs were released by major vendors. Some of these are reasoning models designed to solve complex tasks by decomposing them into smaller steps and applying logical reasoning. In April, OpenAI released o3 and o4-mini, describing o3 as their most powerful reasoning model and o4-mini as a smaller, cost-efficient alternative. They also released non-reasoning models: GPT-4.5 in February and GPT-4.1 in April. Anthropic released Claude 3.7 Sonnet in February, and Google DeepMind introduced Gemini 2.5 Pro in May, both of which are reasoning models. To date, their performance in diagnostic radiology, especially in image interpretation, remains largely unexplored.

This study aimed to evaluate the performance of recent vision-enabled LLMs on the JDRBE, an exam requiring comprehensive expertise in diagnostic radiology. In addition to measuring accuracy, we also assessed the legitimacy of responses based on subjective ratings by radiologists.

## Materials and methods

### Study design

This retrospective study did not directly involve human subjects. All data are devoid of any information that can identify individuals, and are available online to members of Japan Radiological Society (JRS). Model inputs were submitted via application programming interfaces (APIs) of OpenAI, Anthropic, Google Cloud, or Azure AI Foundry, as detailed in later sections. Their privacy policies guarantee that data submitted via their APIs are securely handled and not used for model training. Therefore, Institutional Review Board approval was waived.

### Question dataset

The questions included in our study were entirely derived from the JDRBE, which evaluates comprehensive knowledge of diagnostic radiology. Candidates must complete at least five years of training of radiology to be eligible for the JDRBE.

For the JDRBE 2021 and 2023, we used the same dataset as in our previous report [9]. We additionally prepared the dataset for the 2024 examination following the same method. Briefly, we downloaded the examination papers in the Portable Document Format (PDF) from the member-only section of the JRS website, and extracted text and images using Adobe Acrobat (Adobe, San Jose, CA). For most questions, the extracted images were used as-is. For some questions with inappropriate extracted images (such as those with multiple images superimposed), we used screenshots captured from the PDF files instead. The images were either in PNG or JPEG format, with heights ranging from 134 to 1,708 pixels (mean, 456) and widths from 143 to 1,255 pixels (mean, 482). Figure 1 shows an example of a question processed using this method. Questions from the 2022 examination were excluded because we failed to extract relevant data from the PDF file.

**Figure 1.**
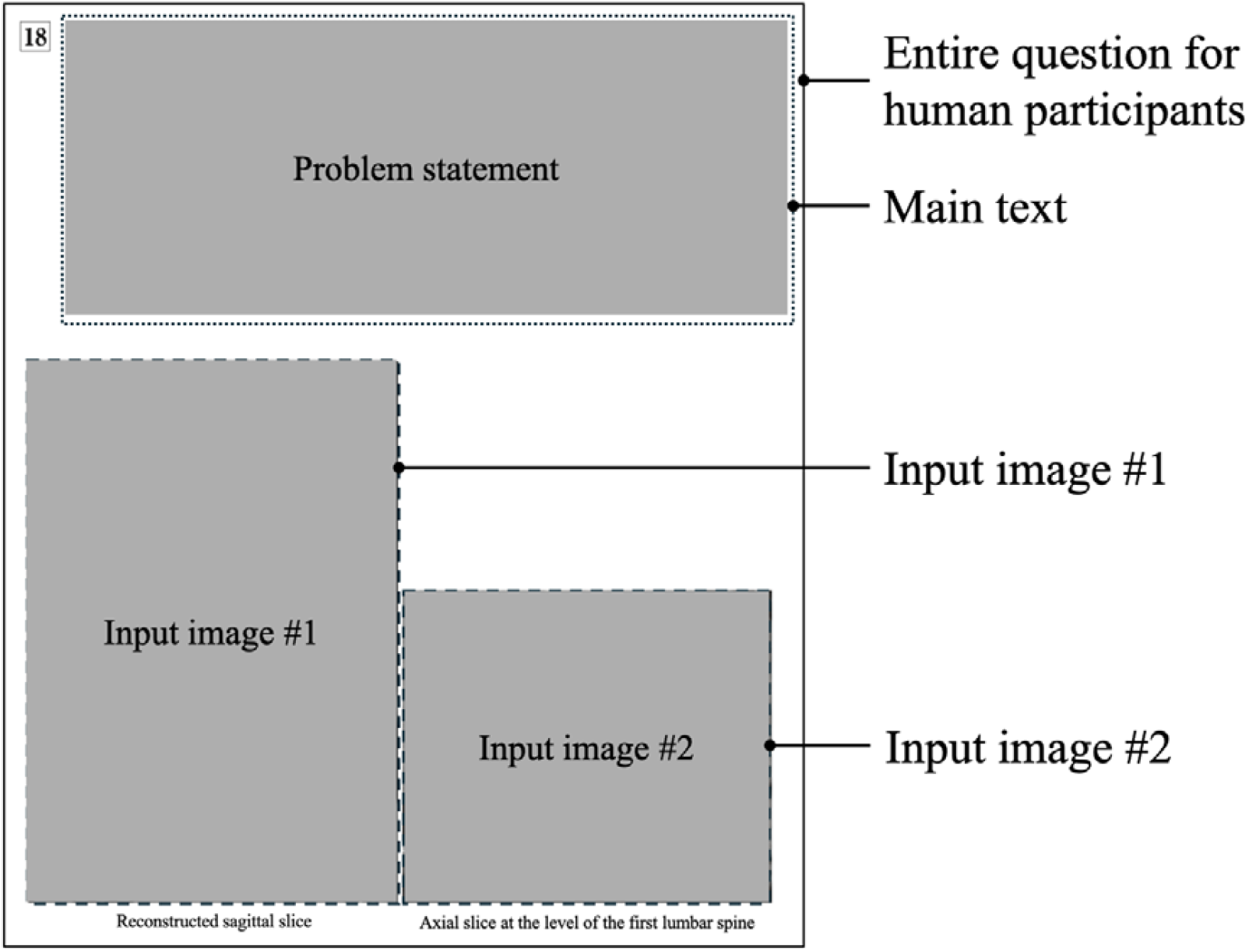
Example of text and image extraction from a question. The main text and input images were extracted and provided to the model, while the question number (“18”) and image captions (“reconstructed sagittal slice” and “axial slice at the level of the first lumbar spine”) were omitted. In this example, the main text states, “A reconstructed sagittal slice and an axial slice at the level of the first lumbar spine are shown.” Due to copyright restrictions, the actual problem statement and images cannot be displayed

Questions without images were excluded; the remaining questions contained one to four images each. All questions had five answer choices. Approximately 90% were single-answer questions, while the remaining 10% were two-answer questions that required choosing both correct answers. The required number of choices was specified in each question statement.

Since there were no officially published answers to the examinations, ground-truth answers were determined through consensus by three or more board-certified diagnostic radiologists. The answers for the 2024 examination were determined by S.M., S.H., and T.Y., with 18, 23, and 30 years of experience in diagnostic radiology, respectively. Questions without unanimous agreement on answers were excluded from the study. Figure 2 illustrates a flow chart detailing the inclusion and exclusion processes for questions.

**Figure 2.**
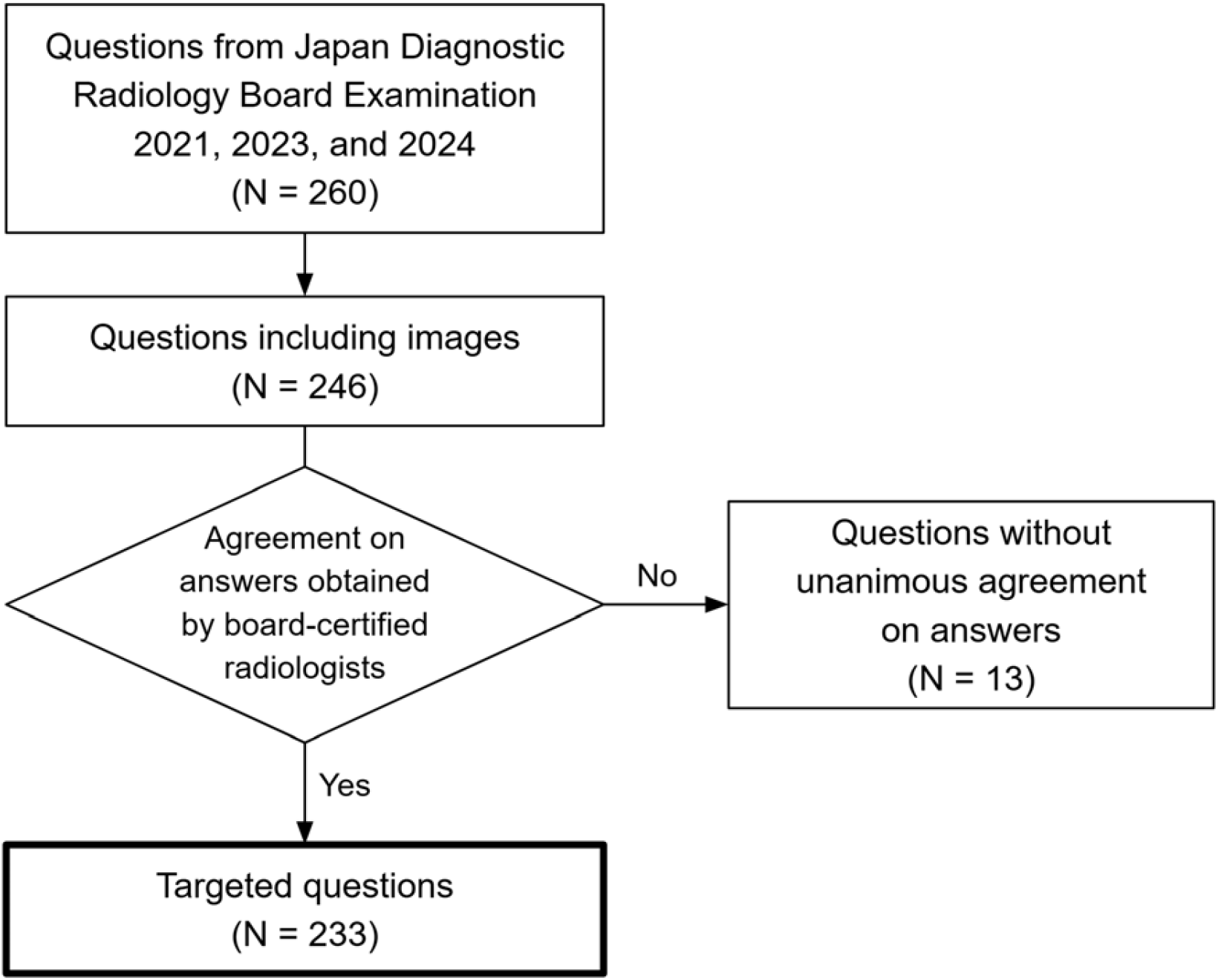
Summary of questions included in this study

### Model evaluations

The following eight models were evaluated: GPT-4 Turbo, GPT-4o, GPT-4.5, GPT-4.1, o3, and o4-mini (all developed by OpenAI, San Francisco, CA); Claude 3.7 Sonnet (Anthropic, San Francisco, CA); and Gemini 2.5 Pro (Google DeepMind, London, UK). Details of the models are described in Table 1. Some of these models are reasoning models designed to solve complex tasks by employing logical reasoning [12–14]. GPT-4 Turbo and GPT-4o were chosen as baselines for comparison with our previous reports; the others are recent models released between February and April 2025. Since the GPT-4 Turbo model used in our previous study was a preview version and has since become unavailable, we used the closest available version in the same model family. All models were tested under two conditions: one using both text and image inputs (hereafter, “vision”) and one using only text inputs (“text-only”).

**Table 1.** Details of tested large language models.

| Model name | Developer | First release date | Version | Knowledge cutoff | Reasoning model |
| --- | --- | --- | --- | --- | --- |
| GPT-4 Turbo | OpenAI | 6 Nov 2023 | *gpt-4-turbo-2024-04-09* | Dec 2023 | No |
| GPT-4o | OpenAI | 13 May 2024 | *gpt-4o-2024-11-20* | Oct 2023 | No |
| GPT-4.5 | OpenAI | 27 Feb 2025 | *gpt-4.5-preview-2025-02-27* | Oct 2023 | No |
| GPT-4.1 | OpenAI | 14 Apr 2025 | *gpt-4.1-2025-04-14* | Jun 2024 | No |
| o3 | OpenAI | 16 Apr 2025 | *o3-2025-04-16* | Jun 2024 | Yes |
| o4-mini | OpenAI | 16 Apr 2025 | *o4-mini-2025-01-31* | Jun 2024 | Yes |
| Claude 3.7 Sonnet | Anthropic | 24 Feb 2025 | *claude-3-7-sonnet-20250219* | Nov 2024 | Yes |
| Gemini 2.5 Pro | Google<br>DeepMind | 28 Mar 2025 | *gemini-2.5-pro-preview-03-25* | Jan 2025 | Yes |

We used the official Anthropic API for Claude 3.7 Sonnet, the Google Cloud API for Gemini 2.5 Pro, and either the OpenAI API or the Azure AI Foundry (Microsoft, Redmond, WA) API for OpenAI models. For Claude 3.7 Sonnet, the required *max_tokens* parameter was set to 4,096; all other parameters for this model and all parameters for the other models were left at their default values. All questions from the examinations were in Japanese, and the textual data were passed to the models without translation. We provided a system prompt similar to the one used in our previous report, as shown in Table 2 (translated into English; please contact the corresponding author for the original Japanese prompts). All experiments were conducted between April 18 and May 1, 2025.

**Table 2.** Prompts used in the experiments.

|  | Prompt (translated into English) |
| --- | --- |
| Vision | You are a radiologist who is about to take the Japan Diagnostic Radiology Board Examination.<br>Below we present a question for the examination and ask you to answer it. Please also briefly explain the thought process that led you to your answer. Even if you are not confident, you will always be forced to select and provide an answer. Please clearly indicate your answer in the format of "ANSWER: x" or "ANSWER: x, y" at the last line. |
| Text-only | You are a radiologist who is about to take the Japan Diagnostic Radiology Board Examination.<br>Below we present a question for the examination and ask you to answer it. Please also briefly explain the thought process that led you to your answer. Note that you will be given only the text of the questions, without any images. Even if you are not confident, you will always be forced to select and provide an answer. Please clearly indicate your answer in the format of "ANSWER: x" or "ANSWER: x, y" at the last line. |

### Legitimacy assessment

To assess the legitimacy of model responses, two diagnostic radiologists with different levels of experience (Y.Y., 2 years; S.M., 18 years, board-certified) independently rated the responses from four models—GPT-4 Turbo, Claude 3.7 Sonnet, o3, and Gemini 2.5 Pro—for all 92 questions from the 2024 examination. A five-point Likert scale (1 = very poor to 5 = excellent) was used to rate each of the 368 responses based on a comprehensive assessment of response quality, including image interpretation, reasoning, and explanation. The responses were presented in randomized order, and the raters were blinded to the model identities.

### Statistical analysis

Differences in performance between the vision and text-only results were analyzed using McNemar’s exact test. For the legitimacy scores, we first applied Friedman’s test, followed by Wilcoxon’s signed-rank test with Holm’s correction for post-hoc pairwise comparisons. Statistical significance was set at P < 0.05. All analyses were conducted using Python (version 3.12.6) with the scipy (version 1.15.2) and statsmodels (version 0.14.4) libraries.

## Results

The dataset comprised 233 questions with 477 images (including 184 CT, 159 MRI, 15 X-ray, and 90 nuclear medicine images). Of these, 210 were single-answer and 23 were two-answer questions. Figure 3 provides an example question, along with summarized responses from the four models rated by the radiologists. The original full responses for this question (translated into English) are available in Supplemental Figure 1.

**Figure 3.**
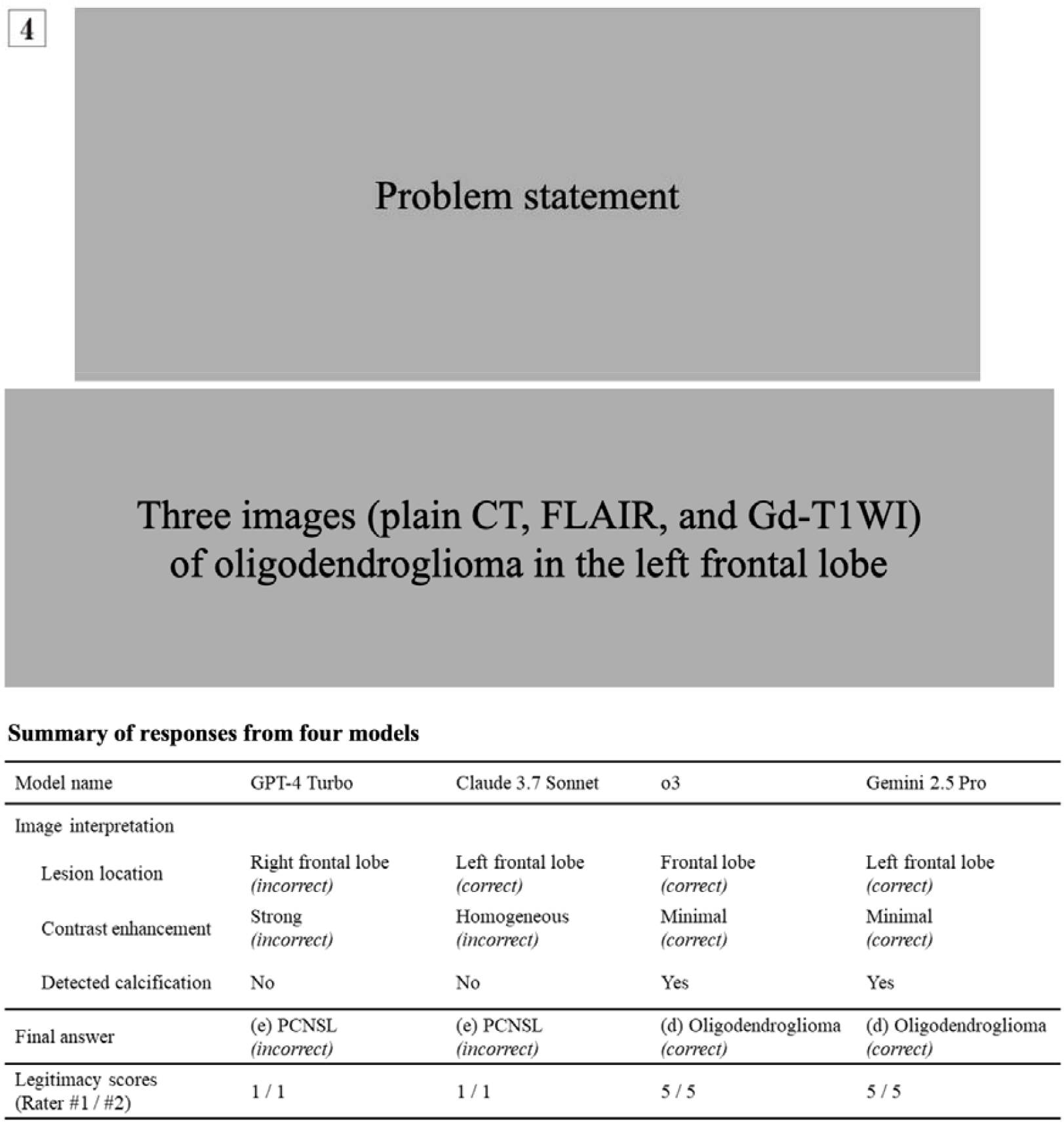
Question 4 from the Japan Diagnostic Radiology Board Examination 2024, representing a clinical scenario of a man in his 30s presented with transient dysphasia. The question asks to identify the most probable diagnosis from the following options: (a) glioblastoma, (b) hemangioblastoma, (c) metastatic brain tumor, (d) oligodendroglioma, and (e) primary central nervous system lymphoma (PCNSL). The correct answer is (d) oligodendroglioma. The figure also includes a summary of responses from four large language models, along with their legitimacy scores rated by diagnostic radiologists. Due to copyright restrictions, the actual problem statement and images cannot be displayed

Table 3 shows the number of correct answers for each model under each condition. The best-performing model was o3 under the vision condition, correctly answering 168 questions (72%). For GPT-4.5 and Gemini 2.5 Pro, significantly higher accuracy was observed in the vision condition compared to the text-only condition. For the other models, no significant difference in accuracy was observed between the two conditions.

**Table 3.** Number of correct responses out of 233 questions, with (vision) and without (text-only) input images.

| Model name | Vision | Text-only | P-value<br>(vision vs. text-only) |
| --- | --- | --- | --- |
| GPT-4 Turbo | 97 (42%) | 102 (44%) | 0.583 |
| GPT-4o | 121 (52%) | 113 (48%) | 0.302 |
| GPT-4.5 | 155 (67%) | 138 (59%) | 0.030 * |
| GPT-4.1 | 141 (61%) | 129 (55%) | 0.134 |
| o3 | 168 (72%) | 156 (67%) | 0.126 |
| o4-mini | 163 (70%) | 151 (65%) | 0.134 |
| Claude 3.7 Sonnet | 127 (55%) | 132 (57%) | 0.590 |
| Gemini 2.5 Pro | 162 (70%) | 137 (59%) | 0.001 * |

\* $P < 0.05$

Figure 4 illustrates the distribution of the legitimacy scores for each rater. Gemini 2.5 Pro received the best scores from both raters (medians, 4 and 5), followed by o3 (4 and 4.5), Claude 3.7 Sonnet (3 and 3), and GPT 4 Turbo (2 and 2). Friedman’s test indicated significant differences among the four models for both raters (P < 0.001). For Rater #1, all six pairwise comparisons showed significant differences after Holm correction: GPT-4 Turbo vs. all others (P < 0.001), Claude 3.7 Sonnet vs. o3 (P = 0.027), Claude 3.7 Sonnet vs. Gemini 2.5 Pro (P < 0.001), and o3 vs. Gemini 2.5 Pro (P = 0.027). For Rater #2, five of the six comparisons were significant: GPT-4 Turbo vs. all others (P < 0.001), Claude 3.7 Sonnet vs. o3 (P = 0.006), and Claude 3.7 Sonnet vs. Gemini 2.5 Pro (P = 0.004). The difference was not significant between o3 and Gemini 2.5 Pro (P = 0.196) for Rater #2.

**Figure 4.**
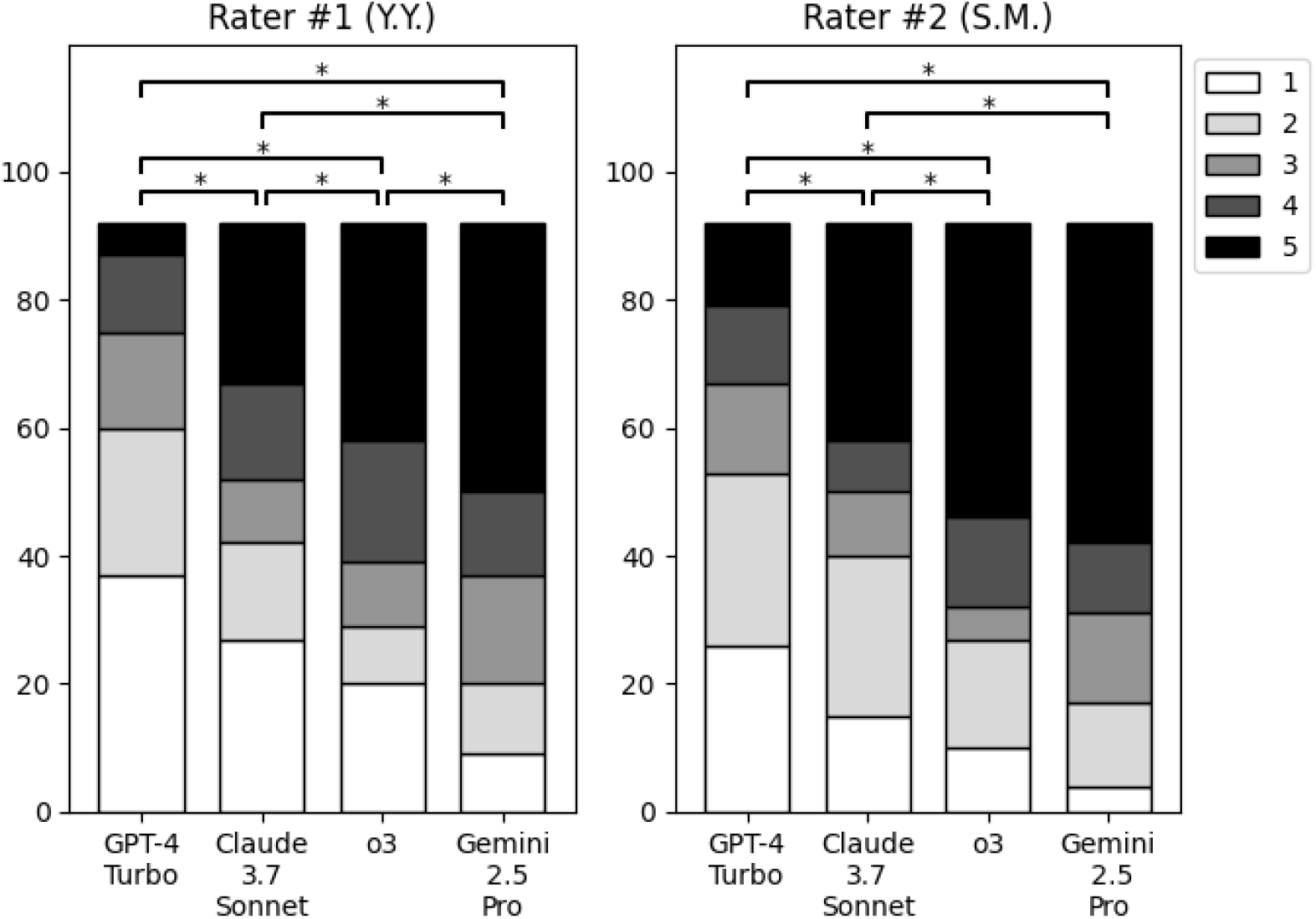
Distribution of legitimacy scores from two raters. * P < 0.05 (after Holm correction)

## Discussion

Since our last report in June 2024, LLMs have significantly improved their accuracy and legitimacy on JDRBE. Both GPT-4 Turbo and GPT-4o were outperformed by all the other models. Particularly, OpenAI’s o3 and Google DeepMind’s Gemini Pro 2.5 achieved a substantial leap in performance, although these models were released no more than 18 months after the debut of GPT-4 Turbo with Vision. To our knowledge, this is the first study that showed that the addition of images achieved a statistically significant accuracy improvement in the JDRBE.

OpenAI’s o3 was the best-performing model under both conditions, achieving an accuracy of 72% with image input. Notably, it correctly answered 67% of the questions even without image input, suggesting its strong medical knowledge and ability to reason the most likely answer based solely on the question text and the provided options. The addition of images did not significantly improve its accuracy, which may indicate that o3 heavily relies on its text-based reasoning capability when choosing an answer. OpenAI explains that reasoning models “think before they answer,” generating a long internal chain of thought before responding to the user [12]. Our results showed that models with reasoning capabilities generally performed better, suggesting that the reasoning ability may be one of the contributing factors in the field of diagnostic radiology.

Gemini 2.5 Pro showed the greatest improvement in accuracy with the addition of images (from 59% to 70%). Furthermore, its legitimacy scores tended to be higher than those of o3, with a statistically significant difference from one of the raters. Although o3 was better in terms of accuracy, these findings suggest that Gemini 2.5 Pro can perform more reasonable medical image interpretation than o3. GPT-4.5 was another model that showed significantly better accuracy with image input.

Figure 3 highlights the ability of o3 and Gemini 2.5 Pro to accurately recognize and describe key image findings. GPT-4 Turbo incorrectly identified the lesion as being in the “right frontal lobe,” reflecting the same left-right confusion reported in our previous study [4]. It selected an incorrect option (malignant lymphoma) and received legitimacy scores of 1 (very poor) from both raters. Claude 3.7 Sonnet successfully identified the lesion location but inaccurately described the lesion as having “homogeneous contrast enhancement,” selecting the same incorrect option (malignant lymphoma) and receiving scores of 1 from both raters. In contrast, both o3 and Gemini 2.5 Pro accurately described the contrast enhancement as minimal and identified small calcification, leading to the correct diagnosis of oligodendroglioma. Both models received legitimacy scores of 5 (excellent) from both raters.

This study has several limitations. First, we could not assess the variability of responses across multiple API calls for the same question. Some reasoning models do not provide a configurable “temperature” parameter, making deterministic outputs unattainable. Second, although reasoning models tended to perform better, we could not conclusively determine whether this was truly owing to their reasoning ability or simply due to increased knowledge. Third, some of the examinations analyzed in this study took place before the knowledge cutoffs of certain LLMs. While access to the exam data is restricted to JRS members, we could not fully exclude the possibility of data leak. Lastly, we used only a single prompt throughout the experiments; alternative prompt strategies might have yielded better performance.

In conclusion, this study assessed the performance of multiple vision-enabled LLMs on the Japan Diagnostic Radiology Board Examination. Recent LLMs, particularly o3 and Gemini 2.5 Pro, demonstrated improved accuracy and legitimacy, reflecting notable advancements in their abilities in diagnostic radiology.

## Data Availability

Examination papers containing all the questions used in this study are available to JRS members via the following URL: https://member.radiology-sys.jp/jrsWebMember/member/specialist/specialist_examination.html. Owing to restricted access to these materials, we are unable to publicly share the questions, the models' responses, or our ground-truth answers.

## Acknowledgments

The Department of Computational Diagnostic Radiology and Preventive Medicine, The University of Tokyo Hospital, is sponsored by HIMEDIC Inc. and Siemens Healthcare K.K. We thank the Japan Radiological Society for granting permission to cite questions from the Japan Diagnostic Radiology Board Examination.

## Declarations

## Ethical approval

This study did not directly involve human subjects. All data used in this study have been anonymized and are devoid of any information that could identify individuals, and these are available online to all JRS members. Therefore, approval from the Institutional Review Board was waived.

## Data Availability

Examination papers containing all the questions used in this study are available to JRS members via the following URL: https://member.radiology-sys.jp/jrsWebMember/member/specialist/specialist_examination.html.

Owing to restricted access to these materials, we are unable to publicly share the questions, the models’ responses, or our ground-truth answers.

