## Supplemental Figure 1 for "Assessing accuracy and legitimacy of multimodal large language models on Japan Diagnostic Radiology Board Examination"

a

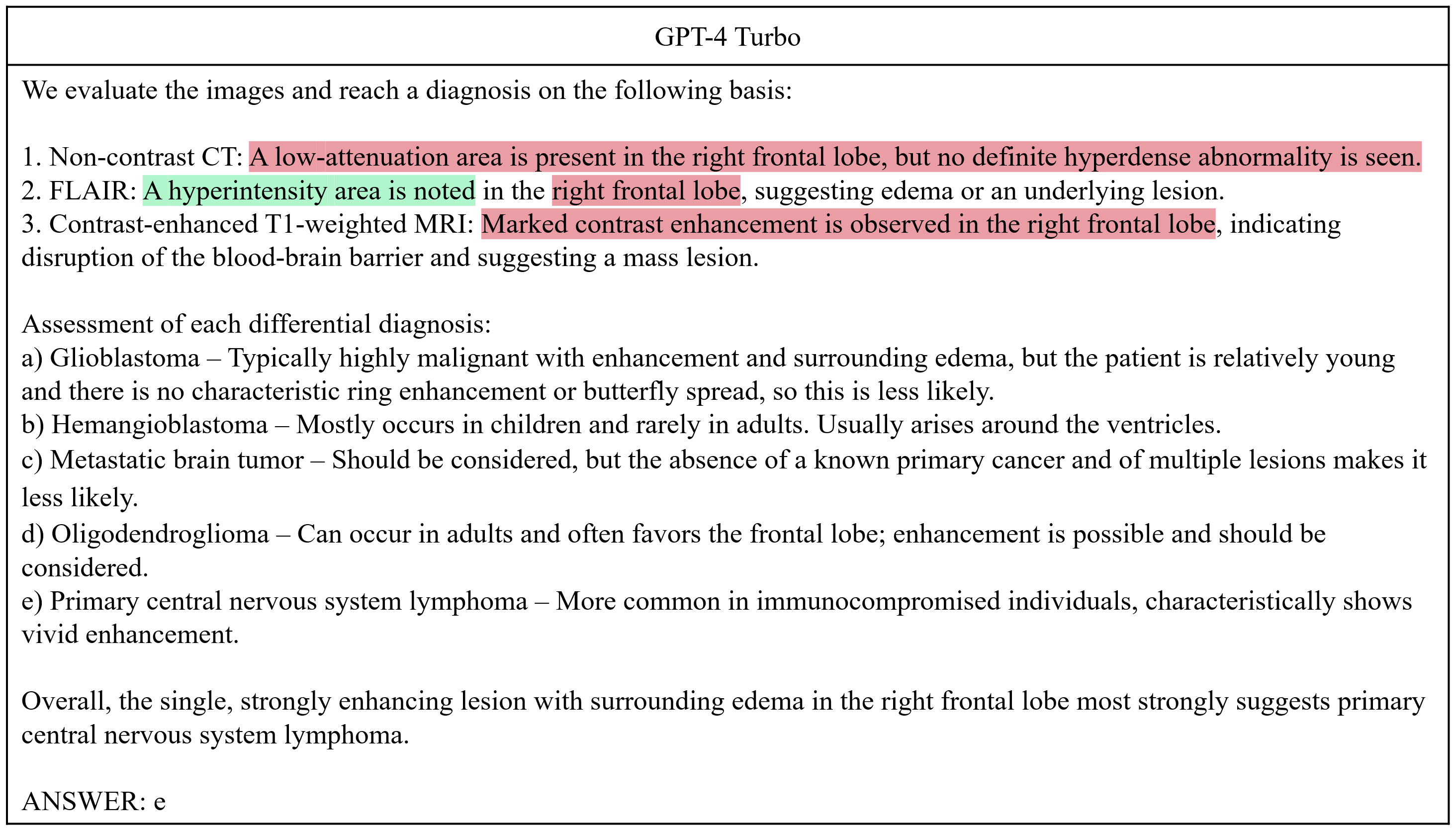

b

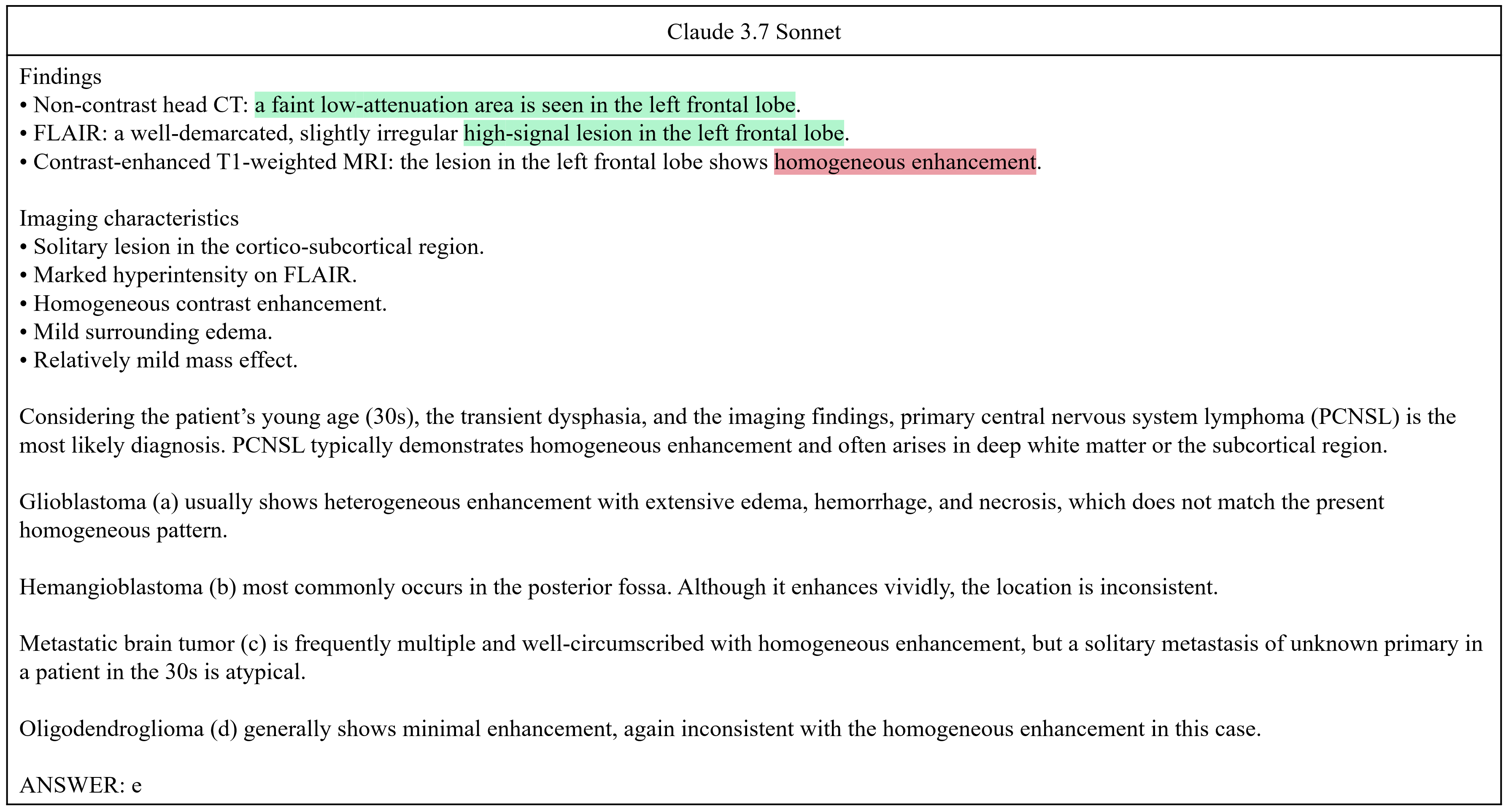

c

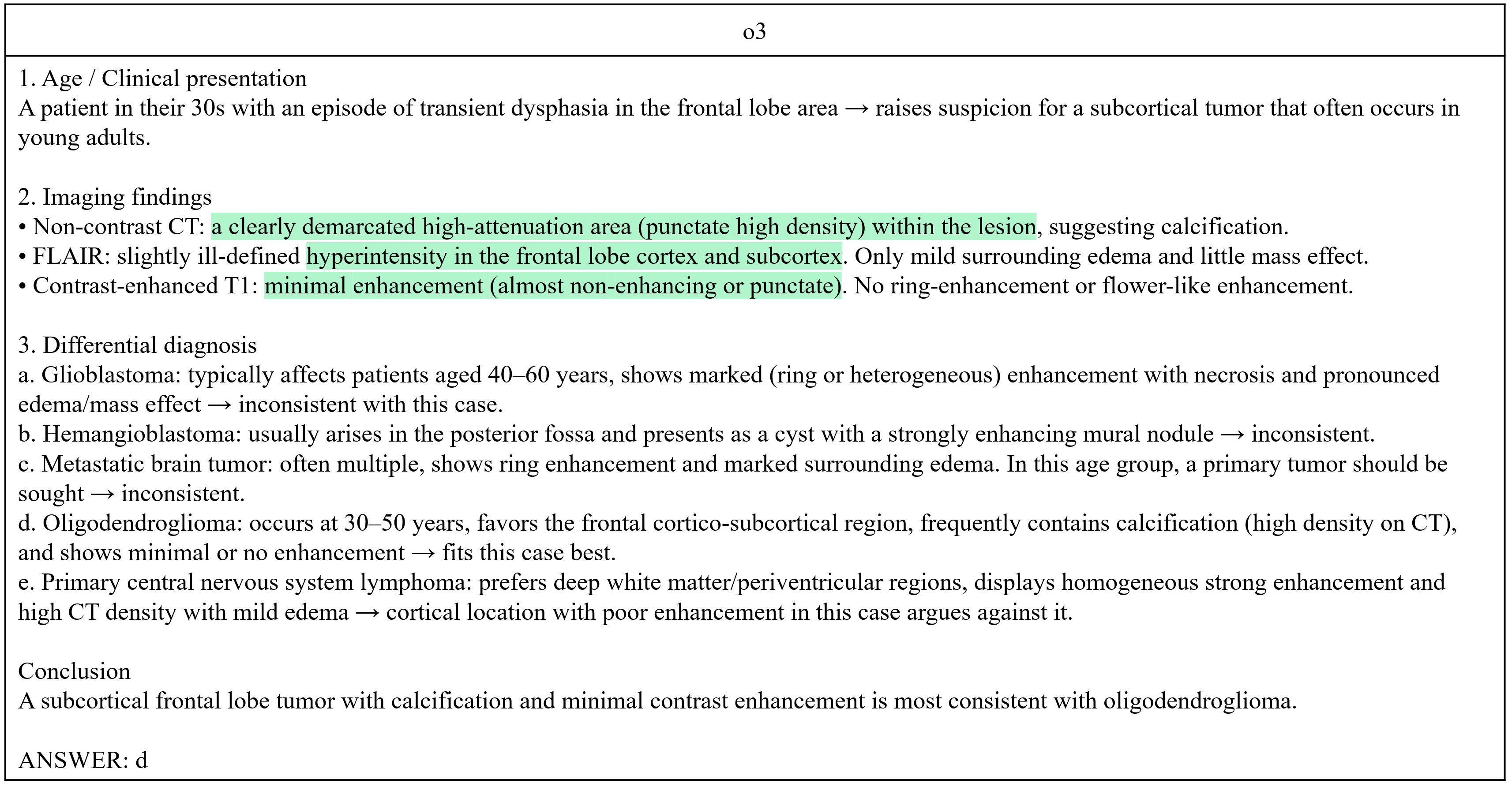

d

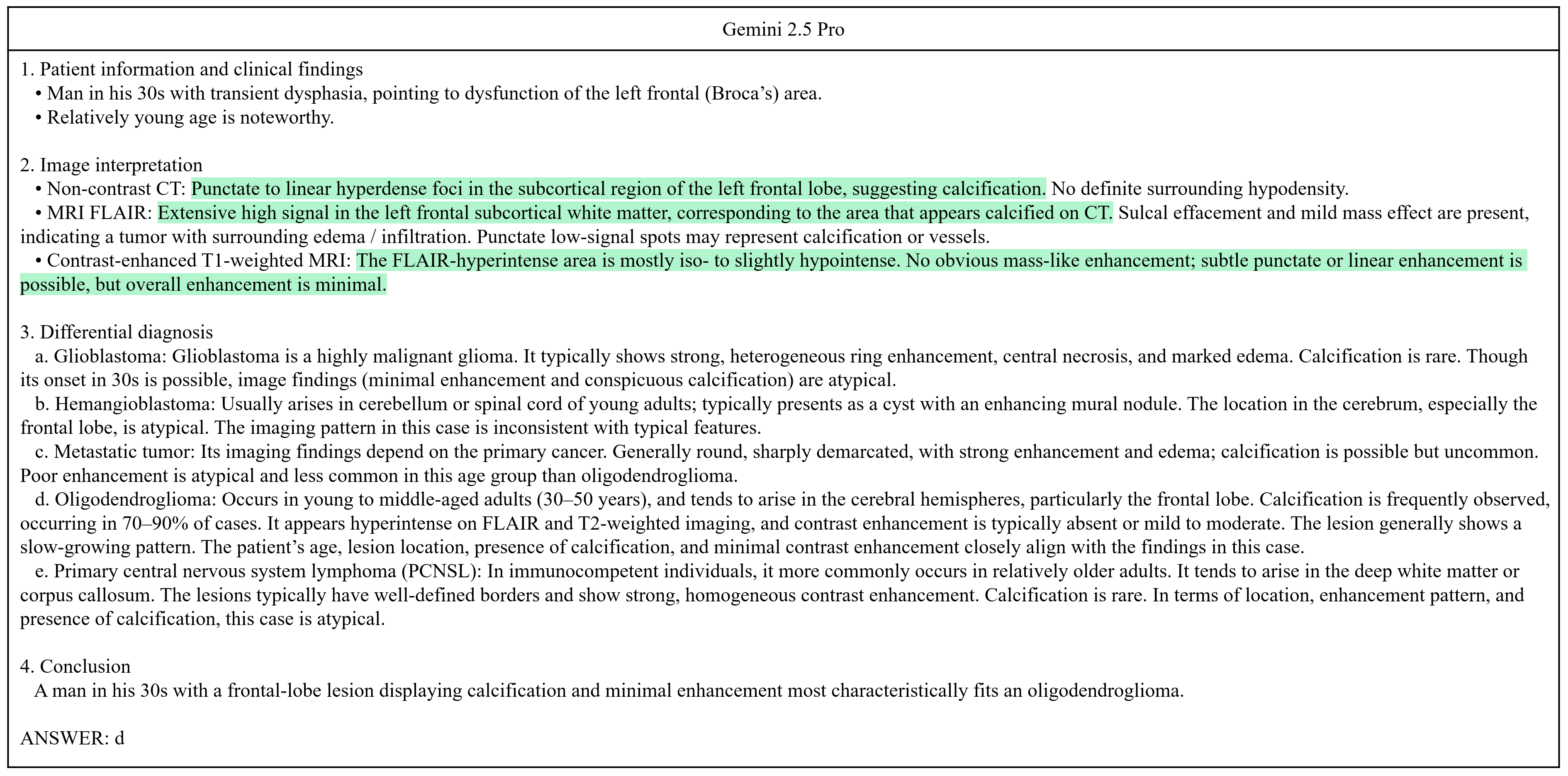

**Supplemental Figure 1** Responses of (a) GPT-4 Turbo, (b) Claude 3.7 Sonnet, (c) o3, and (d) Gemini 2.5 Pro, to the question 4 from the Japan Diagnostic Radiology Board Examination 2024, as described in Figure 3. Text in red and green represents incorrect and correct image interpretation, respectively
